# The neural pathways of change: An fMRI study of the effects of behavioral change suggestions on value-based dietary decision-making

**DOI:** 10.1101/2025.03.03.25323231

**Authors:** Belina Rodrigues, Benjamin Flament, Iraj Khalid, Martine Rampanana, Solene Frileux, Hippolyte Aubertin, Jean-Michel Oppert, Philippe Fossati, Leonie Koban, Christine Poitou, Jean-Yves Rotge, Liane Schmidt

## Abstract

Resolving the ambivalence between immediate cravings and long-term consequences is a key target of communication-based interventions toward behavioral change, such as motivational interviewing. Here, we aimed to understand how this ambivalence influences food preference formation and its neural mechanism. Eighty-five participants with varying body mass indices and food addiction-like symptoms underwent a motivational interviewing session during which they formulated personal reasons for (i.e., change talk) and against (i.e., sustain talk) changing their current eating habits. One week later, they listened to their statements when rating how much they wanted to eat various food items for real at the end of the experiment while their brain activation was measured with functional magnetic resonance imaging. Food choices were healthier when participants had listened to their change talk statements and more based on taste following sustain talk statements. This effect was stronger in participants with overweight and obesity and the more food addiction-like symptoms they displayed in their everyday life eating habits. On the neural level, participants with overweight and obesity also showed stronger functional connectivity between the ventromedial and dorsolateral prefrontal cortex at time of food choice following a change compared to a sustain talk statement. On the contrary, participants with normal to overweight displayed stronger neural craving responses when making their food choices following sustain talk. These findings indicate that shifting between changing and sustaining unhealthy eating habits biases food preference formation and its encoding in neural pathways linked to valuation, cognitive control and craving as a function of weight status.

## Introduction

The transition from good intentions to sustained behavioral changes presents a challenge across diverse domains, including financial planning, environmental sustainability, and public health. One increasingly prevalent manifestation of this intention–behavior gap is unhealthy, addiction-like eating, which affects approximately 20% of individuals in Western societies (1), and contributes to chronic health issues, economic expenditure, and increased mortality and morbidity (2-5). Thus, approaches that address this gap between intention and action are essential for promoting sustainable behavioral change and mitigating long-term adverse outcomes.

Theories of behavioral change propose that a person’s ambivalence to forgo short term and favor long-term rewarding choices is linked to their self-regulation propensities (6), which has led to the development of interventions that target self-regulation via communication (7, 8). One such intervention is motivational interviewing, which has shown promise in substance use rehabilitation (9-12) and in reducing risk-seeking behavior (13, 14). During a motivational interviewing session, personal motivations for change, referred to as change talk, are voiced and contrasted with reasons for keeping current habits, known as sustain talk (15). Counselors guide patients to increase the frequency and strength of change talk, thereby emphasizing and aiming at resolving their ambivalence between reasons for and against change. Despite the evidence for this approach’s effectiveness in instilling behavioral change (13, 14), a key question remains unresolved. One step toward resolving this question is to investigate the neural effects. Notably, how does the brain mediate the ambivalence between changing and sustaining behavioral habits into behavioral outcomes?

Functional brain imaging offers a way to jointly measure the dynamic relationship between behavior and neural activity. Previous work has shown that change talk, compared to sustain talk, increases activation in the dorsolateral prefrontal cortex (dlPFC) among cannabis users (16) and reduces ventral striatum activation in response to alcohol-related cues (17). However, it remains unaddressed how these neural responses relate to changes in decision-making or cravings. Here, we aimed to shed light on the joint acute behavioral and neural effects of mentally transitioning between reasons for changing and sustaining current unhealthy eating habits during dietary decision-making.

We leveraged an approach from decision neuroscience that measures the joint cognitive and neural mechanisms of value-based dietary decision-making, which approximates food preference and its extreme, craving, formation (18, 19). Value-based decision-making has been widely investigated, and results converge on the finding that preference formation across domains recruits the brain’s reward and valuation system formed by the ventromedial prefrontal cortex (vmPFC), ventral striatum, and posterior cingulate cortex (20, 21). Food preference formation requires an integration of several types of information, such as taste, smell, energy state, and a decision maker’s beliefs about these interoceptive and exteroceptive signals and their dieting goals. For instance, the vmPFC has been shown to encode sensitivity to drug and food cravings (22-24). It has also been shown to encode food stimulus values as a function of a decision maker’s beliefs about their energy state (25). Moreover, this hub of the brain’s valuation system has strong positive weights in a recently validated whole brain signature of food and drug craving (26). It acts in synergy with parts of the dlPFC when decision-makers successfully regulated their food cravings or changed their weight status (27-29). Taken together, these findings infer that food preferences and their extreme – cravings, might recruit more than one brain region (30).

Based on these findings, we hypothesized that if motivational interviewing targets a decision-makers ambivalence between changing and sustaining unhealthy eating habits, then listening to change versus sustain talks should: 1) influence self-regulation by how much participants consider the healthiness and tastiness of food during an incentive compatible, dietary decision-making task; and 2) modulate the co-activation of brain regions involved in valuation and cognitive regulation during choice formation. To account for individual differences, we recruited participants with varying body mass indices (BMI) and number of symptoms of food addiction-like behavior. This variability allowed us to explore the contrast effects of change versus sustain talk when behavior needed indeed to be changed because of weight gain and food addiction-like habits. Moreover, we explored how listening to change versus sustain talk influenced responses of an a priori developed Neurobiological Craving Signature (Koban, et al., 2023) for healthy and tasty foods, and whether BMI moderated these neural responses.

## Results

### Effects of change compared to sustain talk on food valuation

To address the first part of our hypotheses, we tested whether change compared to sustain talk affected food valuation and whether this effect differed in participants with normal weight compared to overweight and obesity. Food valuation was measured by ratings of how much participants wanted to eat food stimuli shown to them after having listened to their change and sustain talks, respectively. We refer to these ratings as stimulus value. A mixed-effects analysis of variance found a main effect of type of talk (χ;2 = 5.7,ß = -0.04, SE= 0.01, p = 0.02). Participants wanted to eat food less after listening to change talks than after listening to sustain talk statements (t(79) = -2.4, p = 0.017, Cohen’s d = -0.27, Figure 1a). The model also detected a main effect of BMI group (<25 versus ≥ 25 kg/m^2^) on food stimulus value (χ;2 = 6.2,ß = 0.11, SE= 0.04, p = 0.01), such that participants with a BMI below 25 kg/m^2^ assigned more value to the food stimuli compared to participants with overweight and obesity (t(158) = 3.3, p = 0.001, Cohen’s d = 0.164). As shown in Figure 1b, complementary to the significant categorical main effect of BMI group, food stimulus value ratings also correlated on a continuous scale to BMI (Pearson’s r = -0.23, p = 0.046), which was also observed for reaction times (Pearson’s r = -0.30, p = 0.008) in line with a significant main effect of BMI group on reaction times (χ;2 = 51.1, ß = 0.07, SE= 0.02, p = 0.001) (Figure 1c). In other words, participants with higher BMIs responded faster. BMI group did not interact with the type of talk participants listened to before rating their food wanting (χ;2 = 0.06,ß = 0.004, SE= 0.02, p = 0.81) or how fast they made their decisions (χ;2 = 0.81,ß = -5.5x10e-4, SE= 6.1x10e-4, p = 0.37).

**Figure 1.**
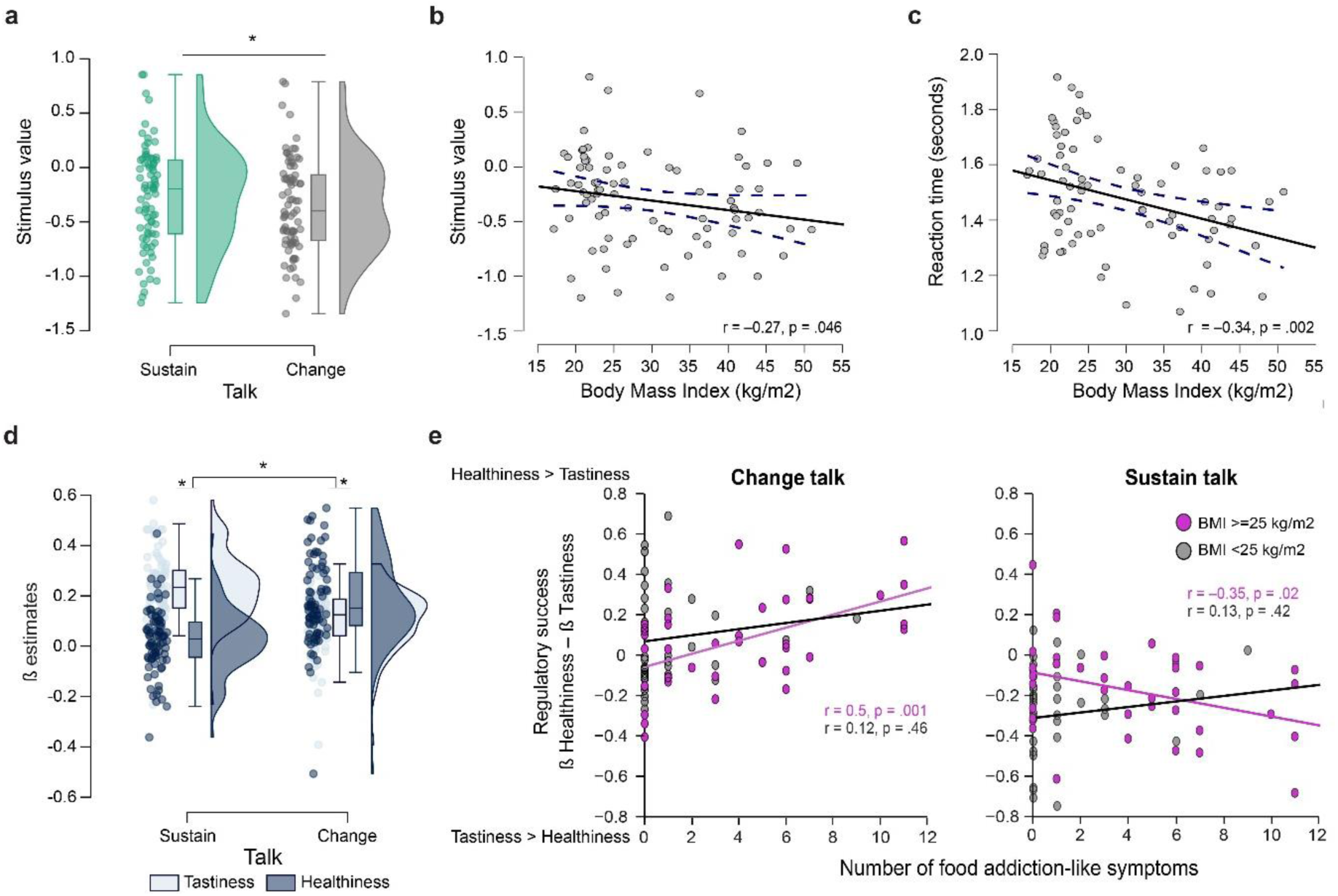
Behavioral results. Boxplots display the 95% confidence intervals of **(a)** the stimulus values (i.e., How much do you want to eat this food? Rated on a 4-point Likert scale) assigned to food during the dietary decision-making task after listening to change and sustain talk statements (n=80 participants). Boxes correspond to the interquartile range from Q1 25^th^ percentile to Q3 75^th^ percentile. The black lines indicate medians and whiskers range from minimum to maximum. The dots correspond to individual participants. Grey boxes correspond to change talk conditions, green boxes to sustain talk conditions. *p < 0.05, paired, two-tailed t-tests within participants. Correlations between body mass index (BMI) and **(b)** stimulus value ratings and **(c)** reaction times. Each dot corresponds to a participant; the blue lines designate 95% confidence intervals. **(d)** Boxplots of beta estimates for healthiness and tastiness in change and sustain talk conditions from a general linear model of stimulus value ratings. **(e)** Correlations of regulatory success scores to number of food addiction-like symptoms in participants with normal BMI (grey dots, n=39, < 25 kg/m^2^) and overweight to obese BMI (pink dots, n=41, ≥ 25 kg/m^2^) following listening to change (left panel) and sustain talk statements (right panel). Regulatory success scores were calculated from the differences in beta estimates shown in panel (**d)** for healthiness minus tastiness effects on stimulus value. Positive scores indicated that the healthiness predicted stimulus values more than the tastiness. Negative scores indicated that the tastiness predicted stimulus value ratings more than the healthiness.

We next tested how listening to change versus sustain talk affected the food preference formation more precisely. Our hypothesis about the cognitive effects assumed that listening to change versus sustain talk influences food valuation by how much a decision-maker considers attributes of food such as its healthiness and tastiness. Our experimental paradigm allowed testing this part of the hypothesis by using a multilevel general linear regression analysis of food stimulus value ratings (i.e., wanting ratings) predicted by individual and self-reported healthiness and tastiness ratings. Note, for each participant the baseline was implicit. The multilevel linear regression modeled how much of the variance in food stimulus value was explained by listening to change relative to sustain talk, and in interaction with a participant’s individual tastiness relative to healthiness ratings of the food stimuli. To this aim, individual beta coefficients for the effects of tastiness and healthiness on food stimulus value in each talk condition were fit into a second level random effect analysis of variance, which detected a significant main effect of food attribute (χ;2 = 16.8, p = 0.001) and an interaction of talk type by attribute (χ;2 = 77.9, p = 0.001). On average, participants considered the tastiness of food more than its healthiness, and even more so after listening to sustain compared to change talk. Importantly, participants changed how much they considered taste over health after listening to their change talks. They now considered the healthiness of food more than its tastiness (Figure 1d).

### Effects of change versus sustain talk and weight status on regulatory success

We next tested our hypothesis that the effects of listening to change relative to sustain talk should be more pronounced in participants, who need to change due to BMI≥25kg/m^2^ or food addiction-like eating habits.

To this aim we calculated for each participant regulatory success scores after each talk condition, which were based on the beta estimates for the effect of healthiness and tastiness on food stimulus value (wanting) ratings. These scores reflected the individual interaction effect, i.e., how much a participant preferred healthy food items over tasty food items after listening to change and after listening to sustain talk statements, respectively. Positive scores meant more regulatory success, in other words, the healthiness of food predicted food wanting more than its tastiness. Negative scores meant that tastiness predicted food wanting more than its healthiness. Participants were grouped into two weight classes, a low BMI group (n=39, BMI < 25 kg/m^2^) and an overweight and obese BMI group (n = 41, BMI ≥ 25 kg/m^2^).

A mixed effects ANOVA of regulatory success scores found a main effect of talk (CT > ST: χ;2 = 17.83, p = 0.001), a non-significant effect of BMI group (low > high BMI: χ;2 = 0.65, p = 0.42) and a significant interaction talk by BMI group (χ;2 = 12.8, p = 0.001). Moreover, the model also found a significant interaction for number of food addiction-like symptoms by talk (χ;2 = 4.13, p = 0.04) and a significant three-way interaction food-addiction score by talk by BMI group (χ;2 = 3.8, p = 0.05).

To unpack these interactions, Figure 1e shows post-hoc correlations between regulatory success scores in change and sustain talk conditions, and as a function of number of food addiction-like symptoms. Participants with a greater number of food addiction-like symptoms showed more regulatory success following change than following sustain talk, which is in line with the interaction food addiction-like symptoms by talk. However, the moderation of regulatory success by food addiction-like symptoms was significant in the obese and overweight BMI group. It was positive for change talk and negative for sustain talk conditions (Pearson’s r = 0.5, p = 0.001 for change talk, and r = –0.35, p = 0.02 for sustain talk), and non-significant in participants with healthy, low BMIs (Pearson’s r = 0.12, p = 0.46 for change talk: Fisher’s r to z = 1.8, p = 0.06, two-tailed and r = 0.13, p = 0.42 for sustain talk: Fisher’s r to z = –2.13, p = 0.03, two tailed). These findings indicated that in participants with higher number of food addiction-like symptoms, who are overweight or obese, listening to change talk statements generated more health-driven food choices, and listening to sustain talk generated more taste-driven food choices. In other words, the effects of the statements were strongest in overweight-to-obese participants with more food addiction-like symptoms.

### Brain imaging results

#### Effects of type of talk and body mass index on value encoding and choice-related brain activation

We next tested our hypothesis about the neural effects of listening to change versus sustain talk during value-based dietary decision-making. We first localized brain activation in correlation to food stimulus value ratings (i.e., how much do you want to eat this food?), and found, in line with the literature, significant activations in regions forming the brain’s valuation system, such as the ventromedial prefrontal cortex (vmPFC), and posterior cingulate cortex (Figure 2a, p_FWE_<0.05, whole brain family wise error corrected). The activation extended into other brain regions such as the inferior frontal gyrus, dorsolateral prefrontal cortex and precuneus (Table 1).

**Figure 2.**
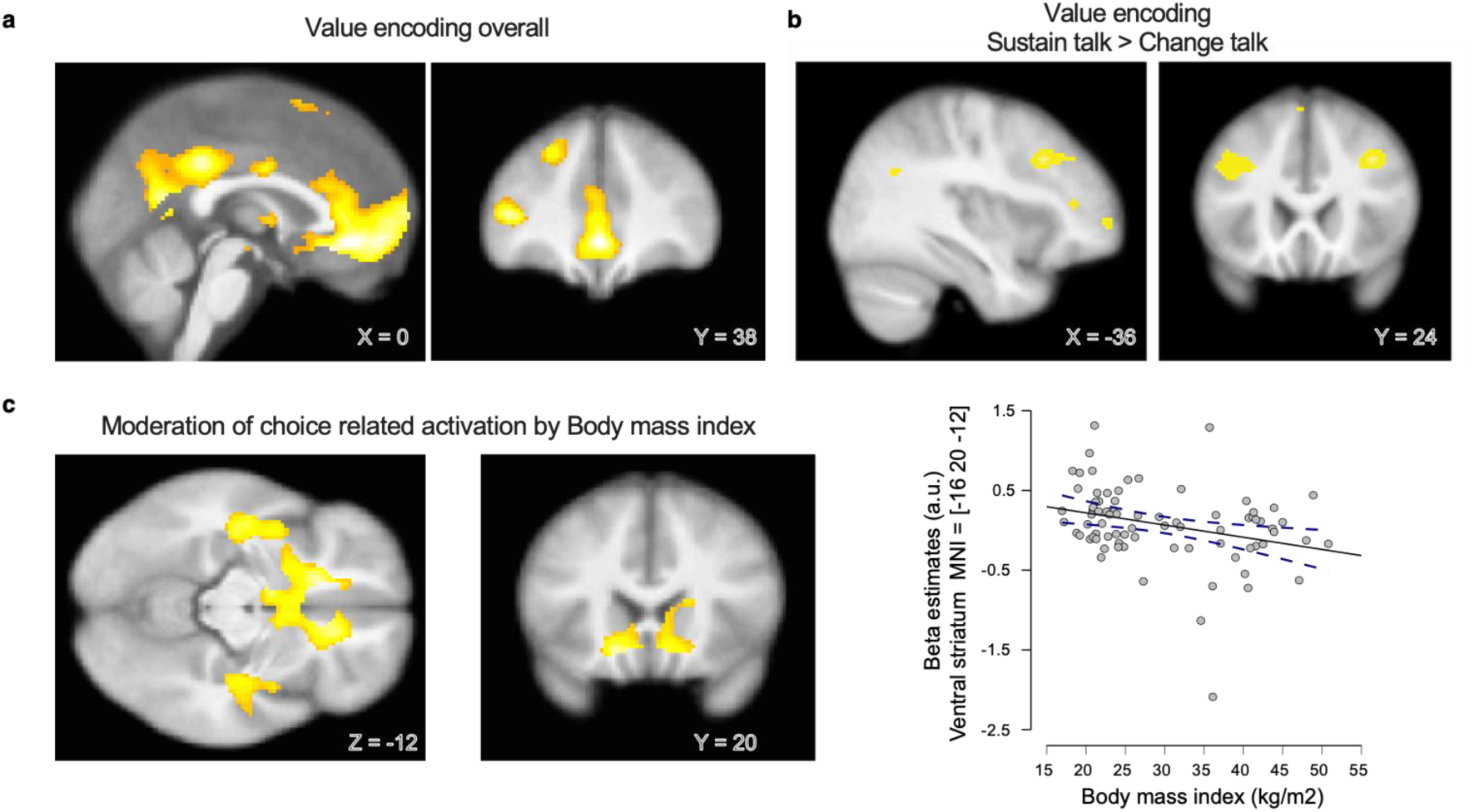
Brain imaging results. Statistical parametric maps (SPMs) show **(a)** correlations to food stimulus value at time of choice (p<0.001 uncorrected for display, survived pFWE<0.05 family wise error whole brain correction, peak and cluster level), and **(b)** how this correlation varied when participants listened to their sustain compared to listening to their change talk (p<0.005 uncorrected, PFWE<0.05 small volume corrected). **(c)** Brain activation at onset choice was negatively moderated by body mass index (p<0.001 uncorrected for display, survived pFWE<0.05 family wise error whole brain correction, peak and cluster level). The scatterplot on the right side displays the moderation across participants (n=80) within a 5 mm radius sphere centered on the peak activation located in the bilateral subgenual anterior cingulate cortex and extending into the ventral striatum MNI= [-16 20 -12]. Significant voxels are displayed at an uncorrected whole brain threshold of p<0.001, and survived pFWE<0.05 family wise error corrections on the cluster level across the whole brain.

**Table 1:** Brain activation correlating to stimulus value at time of choice. (pFWE <0.05, family wise error correction at cluster and peak level)

| Region | BA | Cluster size | X | Y | Z | z-value |
| --- | --- | --- | --- | --- | --- | --- |
| Ventromedial prefrontal cortex |  | 812 | 0 | 38 | -6 | 5.62* |
|  | 32 |  | -2 | 28 | -10 | 5.60 |
|  | 10 |  | -2 | 50 | -6 | 5.49 |
| Posterior cingulate cortex |  | 267 | -8 | -56 | 10 | 5.33 |
| Inferior frontal gyrus |  | 53 | -42 | 40 | 8 | 5.20 |
| Middle frontal gyrus | 9 | 21 | -22 | 34 | 40 | 4.75 |
|  | 6, 8 | 28 | -24 | 10 | 60 | 4.66 |
| Middle temporal gyrus | 39, 40, 22 | 6 | -42 | -70 | 28 | 4.61 |
| Middle frontal gyrus | 8, 6, 9 | 5 | -22 | 28 | 50 | 4.53 |

\* seed region for Psycho-physiological interaction (PPI) analysis, reported in table 4 for the psycho-physiological contrast change versus sustain talk at time of choice onset.

Next, we compared the brain activation in response to food stimulus value ratings at time of food choice following a change and sustain talk and found that the dlPFC correlated stronger to food stimulus values after listening to sustain talk statements compared to food choice trials following change talk (Figure 2b, Table 2, p < 0.001 whole brain uncorrected, small volume correction *p_FWE_<*0.05). These differences in value encoding brain regions between change and sustain talk were not moderated by BMI or food addiction, nor interacted categorically with BMI group (< 25 kg/m^2^ versus ≥ 25kg/m^2^).

**Table 2:** Contrast stimulus value encoding for sustain > change talk at time of choice. (p <0.005 uncorrected, initial whole brain threshold for small volume correction of activations located in the dlPFC)

| Region | BA | Cluster size | X | Y | Z | z-value |
| --- | --- | --- | --- | --- | --- | --- |
| Superior frontal gyrus | 10 | 110 | -26 | 60 | 6 | 3.64 |
| Middle frontal gyrus | 9 | 94 | 36 | 22 | 36 | 3.30 |
|  | 9 | 205 | -36 | 24 | 28 | 3.09* |
|  |  |  | -44 | 20 | 36 | 2.81* |
|  |  | 20 | -36 | 50 | -8 | 3.02 |
| Posterior cingulate cortex | 23 | 59 | 2 | -26 | 30 | 3.00 |

\* significant at $p_{FWE} < 0.05$ small volume corrected at cluster level with a z value of 2.88. Small volume correction was based on a sphere of 10-mm radius centered on the following [x y z] MNI coordinates: [-48 15 24], which were reported by Hare et al. Science 2009 for areas activating stronger in successful self-controllers during dietary decision-making.

We then tested the overall brain activation related to choice onset and found that BMI negatively correlated to the activation of the ventral striatum, subgenual anterior cingulate cortex and bilateral insula (p < 0.001 uncorrected, extend threshold k = 40 corresponding to p_FWE_<0.05 cluster level). That is, participants with higher BMIs displayed lesser activation in these brain regions during choice formation (Figure 2c, Table 3) with no difference between change and sustain talk contexts.

**Table 3:** Brain activation, which at time of choice correlated negatively to body mass index. (p<0.001 uncorrected).

| Region | BA | Cluster size | X | Y | Z | z-value |
| --- | --- | --- | --- | --- | --- | --- |
| STG/Insula | 22 | 312 | 54 | -14 | 2 | 4.37* |
|  |  |  | 36 | -22 | -8 | 3.56* |
| sgACC | 13, 25 | 315 | -16 | 20 | -12 | 3.89* |
| Ventral striatum |  |  | -8 | -2 | -12 | 3.69* |
|  |  |  | 4 | 8 | -12 | 3.39* |
| ACC/vmPFC |  | 136 | 12 | 30 | -10 | 3.79 |
| Postcentral gyrus | 5 | 131 | 18 | -42 | 52 | 3.71 |
|  | 31 |  | 6 | -32 | 46 | 3.32 |
| Insula |  | 88 | -42 | -14 | -14 | 3.68 |
| Culmen/PAG |  | 51 | -4 | -42 | -12 | 3.52 |

\* clusters, which form $k \geq 312$ voxels and survive $p_{FWE} < 0.05$ , family wise error correction using a voxel-wise threshold of $p < 0.001$ .

These findings replicated that food wanting was encoded within the brain’s brain valuation system. They further showed that BMI was moderating the activation of subcortical regions of the brain’s valuation system at time of choice formation, and regions linked to the brain’s cognitive regulation system encoded food stimulus values more after listening to sustain talk statements than listening to change talk statements.

#### Effects of type of talk and BMI on functional connectivity of the vmPFC at onset food choice

To test our second hypothesis that proposed that brain activation within valuation and cognitive regulation systems should vary as a function of both type of talk and BMI, we conducted a psychophysiological interaction analysis (PPI). In more detail, the PPI tested (1) how much choice onset activation of a seed region located in the vmPFC covaried with the rest of the brain after listening to change compared to listening to sustain talk, and (2) whether this covariance was different between participants with BMI<25 kg/m^2^ and participants with overweight and obese BMIs (≥25 kg/m^2^).

The PPI detected stronger functional connectivity at time of choice following a change versus a sustain talk between the vmPFC seed region and voxels located within the dlPFC in the participants with overweight and obesity group compared to the normal weight group. The activation spanned across the right superior frontal to the middle frontal gyrus and left anterior cingulate cortex (p_FWE_ < 0.05 cluster level, Figure 3a, Table 4). Other prominent PPI activation that was stronger in participants with overweight and obesity compared to normal weight participants was observed in the primary and premotor cortex (Table 4).

**Figure 3:**
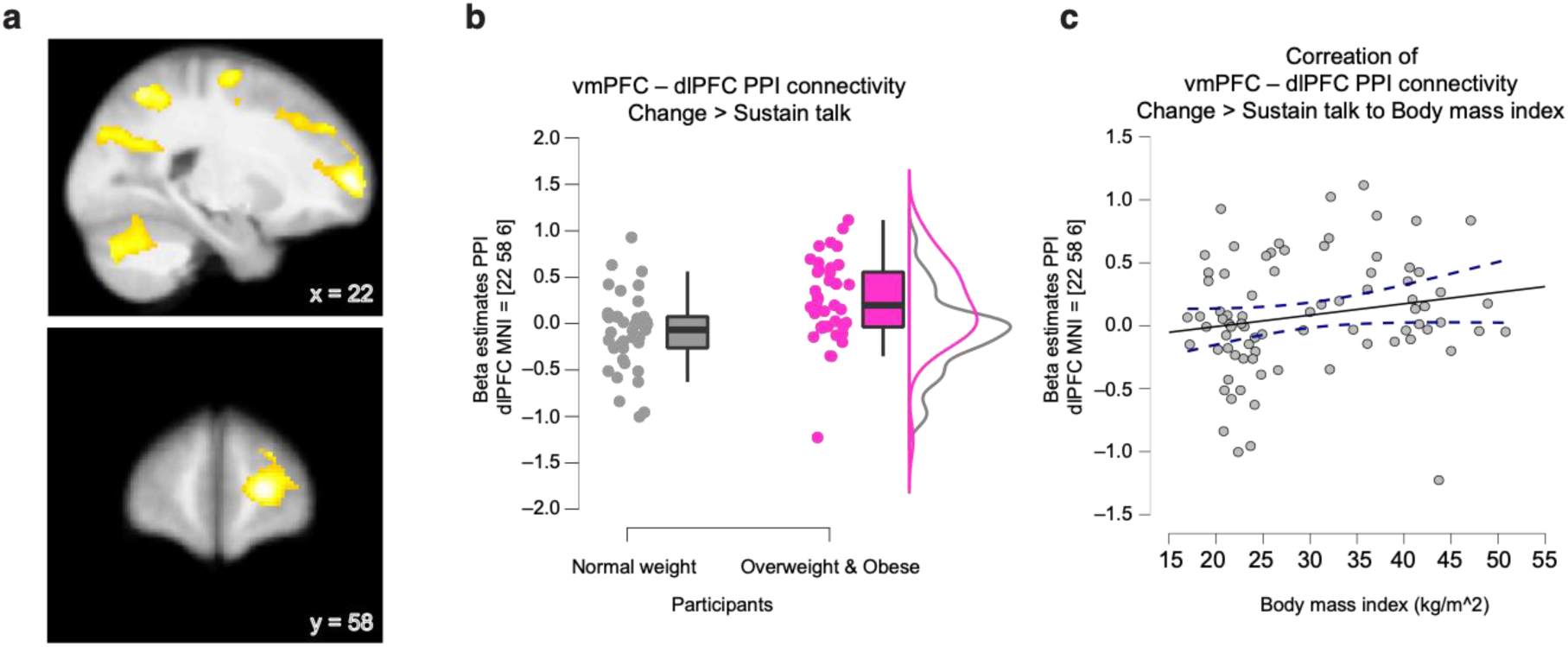
Psycho-physiological interaction (PPI) results. **(a)** SPMs display significant voxels in yellow and orange, with PPI connectivity to the vmPFC seed region that covaried stronger in participants with obesity and overweight than in participants with normal weight after listening to change versus sustain talk. **(b)** Boxplots show 95% CI for dlPFC (MNI=[22 58 6]) to vmPFC – dlPFC covariance for choice onset after listening to change versus sustain talk in participants with normal weight (BMI < 25 kg/m^2^) and in participants with overweight or obesity (BMI ≥ 25 kg/m^2^). Boxes correspond to the interquartile range from Q1 25^th^ percentile to Q3 75^th^ percentile. The black lines indicate medians and whiskers range from minimum to maximum. The dots correspond to individual participants. **(c)** Correlation of body mass index to dlPFC – vmPFC PPI connectivity. Each dot corresponds to a participant. Dotted lines indicate 95% CIs. Note, boxplots shown in **(b)** and scatterplots shown in **(c)** are complementary to the SPMs shown in **(a)** and display the same result in three different ways.

**Table 4:** Contrast participants with overweight and obesity > normal weight participants in PPI related brain activation. (pFWE<0.05 at voxel forming initial threshold of p<0.005 extent threshold k = 1521 voxel)

| Region | BA | Cluster size | X | Y | Z | z-value |
| --- | --- | --- | --- | --- | --- | --- |
| PCC – ACC | 31, 24 | 27381 | –8 | –18 | 48 | 4.31 |
| Fusiform gyrus |  |  | –44 | –36 | –16 | 4.30 |
| SFG/MFG* | 10 |  | 22 | 58 | 6 | 4.28 |
| Precentral gyrus |  | 1521 | 58 | –14 | 40 | 3.62 |
| Primary motor | 4 |  | 46 | –14 | 54 | 3.59 |
| SMA/premotor | 6 |  | 32 | –8 | 62 | 3.53 |

\* used as a MNI coordinate for beta estimate extraction displayed in Figure 3b.

#### Effects of change versus sustain talk on craving-related brain responses

Finally, we tested the hypothesis that food preference formation after listening to change versus sustain talks altered activity in craving-related brain areas. To this aim we tested responses of the previously validated Neurobiological Craving Signature (NCS), which measures activity patterns across the whole brain to predict food craving (i.e., food wanting ratings) (Koban et al. 2023). More specifically, we calculated for each participant the dot products between the NCS (shown in Figure 3a) and the parametric moderation of brain activation at time of choice by stimulus values assigned to tasty food (i.e., reflecting both healthy and unhealthy tasty food wanting) and by stimulus values assigned to healthy food (i.e., both tasty and untasty healthy food wanting) following change and sustain talks, respectively.

We found that the Neurobiological Craving Signature (NCS) estimates were significantly stronger for tasty food stimulus values (i.e., wanting) than for healthy food stimulus values (i.e., wanting) after sustain talk statements. This effect was indicated by a significant interaction type of talk by type of food (χ;2 = 4.77, p = 0.03, Figure 4b) specifically in the participants with normal to overweight. The interaction was driven by a significant difference in NCS estimates of tasty versus healthy food wanting (t(74) = 2.3, p = 0.024, Cohen’s d = 0.26, two-tailed, paired t-test). No significant interactions between type of talk and type of food wanting on NCS responses were observed in participants with obesity.

**Figure 4.**
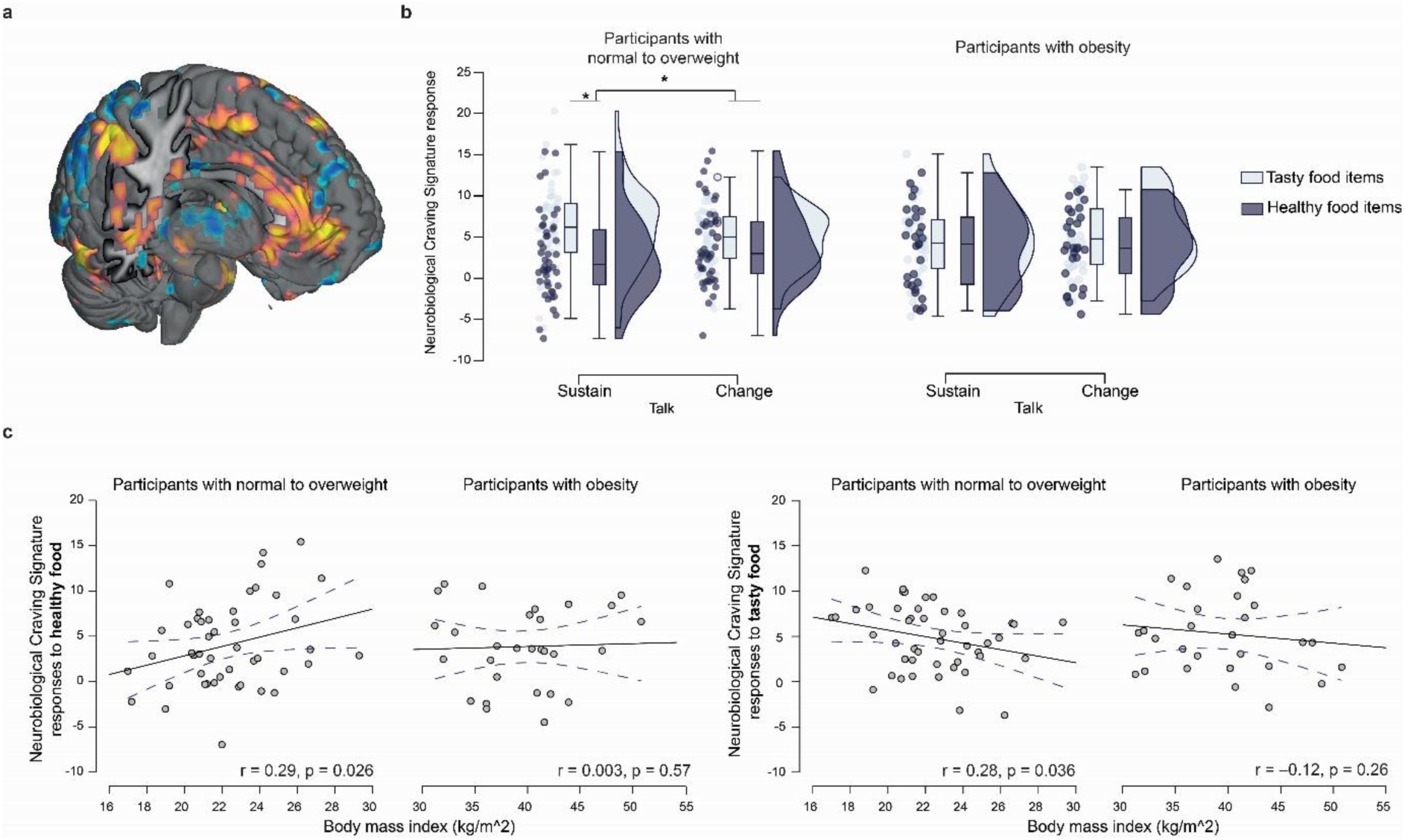
Neurobiological Craving Signature responses following change and sustain talks in participants with normal, overweight and obesity. **(a)** Pattern of the Neurobiological Craving Signature with voxels in yellow displaying positive weights, and in blue displaying negative weights to predict food and drug cravings (at p<0.005 uncorrected for display). **(b)** Interaction type of talk by type of food in normal to overweight participants and participants with obesity. The boxplots display 95% CI for the Neurobiological Craving Signature response to tasty and healthy food stimulus values (wanting). Error bars correspond to standard errors. Horizontal bars indicate median values. Individual points correspond to participants. **(c)** Correlations between body mass indices and the neurobiological craving signature response to healthy and tasty food wanting following listening to change talk statements in participants with normal to overweight and in participants with obesity. Dots correspond to participants; blue dotted lines indicate 95% CIs.

Moreover, as shown in Figure 4c when listening to change talk and across participants with normal to overweight those, with a higher BMI displayed stronger NCS estimates of healthy food wanting (Pearson’s r = 0.29, p = 0.026, n=46, one-sided) and lower NCS estimates of tasty food wanting (Pearson’s r = –0.27, p = 0.036, n=46, one-sided). This moderation of brain activity estimates of tasty and healthy food wanting/craving by BMI was non-significant in participants with obesity. These results suggest that participants within a weight range from normal to overweight showed change talk congruent food craving related brain responses that varied with their BMI. This covariance between change-talk congruent NCS responses and BMI was lacking in the participants with obesity.

## Discussion

Many people are torn between their unhealthy eating habits and their desire to improve health. This study aimed to map this ambivalence between changing versus sustaining unhealthy eating habits — an approach central to motivational interviewing — onto cognitive processes such as self-regulation propensities and their underlying neural mechanisms. To achieve this, we employed value-based dietary decision-making as a model and adopted a theory-driven framework, focusing on BOLD activations in two key brain regions: the ventromedial prefrontal cortex (vmPFC), part of the brain’s valuation system, and the dorsolateral prefrontal cortex (dlPFC), associated with cognitive regulation. Our findings revealed that listening to change talk compared to sustain talk influenced participants’ willingness to forgo the tastiness of food in favor of its healthiness. Among participants with overweight or obesity, regulatory success—measured by their ability to prioritize healthiness—was greater after listening to change talk, especially in those exhibiting more food addiction-like symptoms. At the neural level, we observed that the participants with overweight or obesity displayed stronger PPI connectivity between the value-encoding vmPFC seed region and the dlPFC at the time of food choice following change talk relative to sustain talk. In contrast, responses of the Neurobiological Craving Signature (NCS) to tasty and healthy food wanting were stronger after change talk statements in normal-to-overweight participants.

Our finding of the interaction between the type of talk and type of food attribute (e.g. healthiness, tastiness) is in line with what is known about the how attentional focus can affect value-based dietary decision-making. For example, increased attention toward the tastiness of food has been shown to lead to an increased preference for tasty food. On the contrary, attentional focus on the healthiness shifts preferences toward healthier food options (31, 32). Our findings add a novel perspective on this effect of attentional focus, by showing that self-generated statements rather than explicit instructions can modulate food preferences. This novel finding opens the window into the neurocognitive mechanisms of communication-based therapies such as motivational interviewing for promoting healthier dieting.

We defined the observed propensity to consider the healthiness of food more than the tastiness as regulatory success and found that both BMI and food addiction-like symptoms moderated this propensity. Food addiction was measured by the Yale Food Addiction Scale (33), which posits that processed, highly palatable, and high fat foods can trigger behavioral habits that correspond to addiction-like behaviors defined by DSM-V criteria for substance abuse disorders and addiction-like diseases. Although food addiction is still a debated topic (see reference 34), our findings suggest that listening to change talk statements was more effective in generating healthier food choices in those participants who are likely more aware of the consequences of overweight and obesity and, possibly also because they are already experiencing some of these consequences by presenting addiction-like eating behavior. This conclusion is supported by evidence that individuals with more severe conditions are more responsive to communication-based interventions that foster behavioral change (35).

Given the behavioral differences detected between change and sustain talk conditions, and across a broad range of BMI and food-addiction-like behavioral symptoms, we next asked how these effects were underpinned by brain activation measured with functional magnetic resonance imaging during value-based dietary decision-making. Our finding that brain regions forming the brain’s reward and valuation system, such as the vmPFC and ventral striatum (21) encoded food value less strongly in participants with higher BMI is in line with past work on the links between weight and reward-related brain activation. Notably, this line of research has shown attenuated striatal activation in response to the receipt of palatable foods after weight gain (36), and lower resting state functional connectivity between the vmPFC and ventral striatum in participants with obesity (28). Our finding lines up with this literature by indicating that weight status is linked to the intrinsic functional sensitivity of the brain’s reward and valuation system during food choice formation.

When comparing food stimulus value encoding in the brain between the two talk conditions, we found that the dlPFC correlated more strongly to food stimulus value after listening to sustain talk than after listening to change talk statements. This finding stands at odds with the literature on the self-control of dietary decision-making. Previous findings in this field have shown that food value encoding in the dlPFC during dietary decision-making is a neural marker for more health-driven food choices (27, 31, 37, 38). Differences in study goals, design, and population might explain these odds.

In this study we used the participants’ real reasons for and against changing their eating habits to test their effects on incentive compatible dietary choices. It is possible that hearing their voices, potentially galvanized by the intervention, might have reframed sustain talk into change talk. For example, after hearing a sustain talk statement like: "It is difficult to find the time to cook healthy meals," participants may have considered that while that remained true, they could still try to change this habit. This implies a more implicit process of reframing, which requires more attentional resources and potentially an activation of the dlPFC as shown by previous work on framing effects of long- and short-term consequences of economic choices (39). The change talk statements provide an explicit reason with clear and direct future consequences that may make the decision to reject or want a given food item less cognitively taxing. In contrast, sustain talk may have masked the future consequences of maintaining behavior, thereby increasing cognitive demand and engaging more the dlPFC. Although all participants rated at the end of the fMRI session each statement to be a change and sustain talk, and a reframing was not reflected in the observed food choices, we cannot completely rule out how the participants implicitly conceived their change and sustain talk statements during fMRI.

To some extent the psycho-physiological interaction results can offer a novel perspective on this question. Participants with overweight and obesity displayed stronger functional connectivity after listening to change relative to sustain talk between the vmPFC and the dlPFC at time of food choice formation. Given the prominent role of the dlPFC during self-regulation and attentional resource allocation, its enhanced connectivity to the vmPFC during food choice formation may be compensatory for reaching similar levels of regulatory success as observed in participants with healthy weight. Taken together, we call for more future work to further disentangle the contribution of attentional resource allocation and implicit changes in dieting goals during the decision-making process and following self-suggestions or similar ambivalent mindsets.

Lastly, we have looked at how food preference formation following change and sustain talk statements was underpinned by whole brain-patterns predictive of food craving, beyond the two regions of interest. We combined a recently validated Neurobiological Craving Signature (NCS) with the observed brain activation in correlation to food stimulus value, obtaining a brain-activity based estimate of food wanting/craving. Following change talk statements, this brain activity-based estimate of food wanting increased for healthy food and decreased for tasty food the higher the participant’s BMI. Importantly, these links were only observed within a normal to overweight BMI range. Strikingly, no moderation between NCS responses to healthy and tasty food wanting after change talk were observed in participants with obesity. It is possible that obesity affects food craving related neurobiological responses, which may potentially associate with functional perturbations in the hormonal drivers of food behavior such as leptin (40). Alternatively, weight status may be linked to the recruitment of different neural systems during dietary decision-making following change talk. For example, participants with obesity displayed stronger connectivity between valuation and cognitive control regions, whereas normal weight and overweight participants displayed NCS responses that were sensitive to BMI, type of talk and food. We call for more work to investigate the causality of these weight status moderations of dietary decision-making after change and sustain self-suggestions by for example, testing drastic weight loss interventions in participants with obesity such as bariatric surgery or GLP-1 agonist treatment.

In conclusion, this study investigated how different types of self-generated statements for and against changing eating habits influence dietary decision-making in a heterogeneous participant sample in terms of BMI, and food-addiction-like symptoms. This heterogeneity allowed the exploration of sources for inter-individual differences in the underlying cognitive and neural mechanisms of mentally shifting between change and sustain talks during value-based dietary decision-making. By demonstrating that change talk can enhance regulatory success and influence neural connectivity between the vmPFC and dlPFC, particularly in individuals with higher BMIs and food addiction-like symptoms, this research underscores the importance of tailored interventions in promoting healthier eating habits. Furthermore, the ecological approach of utilizing participants’ self-generated statements adds to the study’s applicability to the real life outside the laboratory. It provides valuable insights into the complexity of behavior change processes and contributes to the growing body of literature on dietary self-regulation but also extends the relevance of motivational interviewing to diverse populations, and its potential for fostering healthier lifestyles.

## Material and methods

### Ethical considerations

The study was approved by the local ethics committees (CPP19.12.05 and CPP20.11.24.38405) and conducted in accordance with the Declaration of Helsinki. All participants provided written and informed consent.

It was pre-registered on the clinical trials website under the following link (https://clinicaltrials.gov/study/NCT05101863). The study deviates from the pre-registered protocol in the following ways:

1. We explored how change and sustain talk influenced neural craving responses to food wanting by using a validated neurobiological craving signature (Koban et al., 2023). This explorative analysis was not initially pre-registered.
2. Due to challenging recruitment of participant with obesity within a clinical setting and a dismantling of the MRI scanner mid-run we could not meet a total balanced sample size of 30 participants without obesity, 30 participants with obesity and 30 participants with obesity and food addiction. Therefore, we conducted categorial group comparisons between participants with normal weight (n = 39), with overweight (n = 7) and with obesity (n = 34), and continuous analyses across the total BMI and Yale Food Addiction Scale (YFAS) ranges.
3. Two participants self-reported normal BMIs above 18.5 kg/m^2^ at the time of recruitment but then a bioimpedance BMI measure of 17 kg/m^2^ at the time of the motivational interviewing session. However, their body fat was in the healthy range (15% and 17%). We therefore did not exclude these participants from analyses, to also prevent unnecessary reduction of the sample sizes.

### Participants

A total of 85 participants were recruited via public advertisement and in collaboration with the Nutrition department at Pitié-Salpêtrière University hospital in Paris. Inclusion criteria for all participants were: age between 18 and 70 years, right-handedness, normal to corrected-to-normal vision. Non-inclusion criteria were history of substance abuse, neurological or psychiatric disorder, and MRI contraindications.

Nine participants were excluded from the analysis due to the following criteria: missing brain imaging data due to MRI scanner breakdowns (n=3), discovery of an fMRI contraindication on the day of the MRI visit (n=2), and head movement (n=4). Thus, a total sample of n = 80 participants were included for behavioral analyses and n = 76 for brain imaging analyses. Among the 80 participants in the behavioral analyses, n = 39 were included in the normal weight group (BMI<25kg/m^2^), while n = 41 were in the group BMI≥25kg/m^2^. For the brain imaging analysis, among the 76 participants, n = 39 were in the normal weight group (BMI<25kg/m^2^), and n = 37 in the group BMI≥25kg/m^2^.

### Experimental procedure

The experimental design involved two visits that took place one week apart.

#### Visit 1

This visit was scheduled on a regular weekday (avoiding weekends and holidays, such as Christmas) to minimize seasonal variations in dietary habits, such as changes in eating patterns on weekends and holidays. Before starting the motivational interviewing session participants reported sociodemographic variables, and filled out a five-question screening tool, the SCOFF questionnaire (41), to rule out the existence of eating disorders. This data is reported in Table 5. Participants further underwent a 24h dietary recall from the previous day’s food intake according to the manual adaptation of the Automated Multiple-Pass Method (42). The 24h dietary recall was used to measure diet quality reflected by the healthy eating index (HEI) following Krebs-Smith et al., 2018 (43) also reported in Table 5.

**Table 5.** Sociodemographic, anthropometric and questionnaire data (n=80)

|  |  |
| --- | --- |
| Mean years of age (SEM) | 35.1 (1.5) years |
| N women (%) | 63 (79%) |
| Mean years of education after high school (SEM) | 3.7 (0.2) years |
| N participants with Professional training (%) | 1 (1.3%) |
| N participants with high school graduates (%) | 6 (7.5%) |
| N participants with at least 2 years of college (%) | 18 (22.5%) |
| N participants with more than 2 years of college (%) | 55 (68.8%) |
| **Body composition** |  |
| Mean BMI (SEM) | 29.9 (1.1) kg/m<sup>2</sup> |
| Range of BMI | 17.0 – 50.8 kg/m<sup>2</sup> |
| **N participants within BMI categories (%)** |  |
| <18.5 | 3 (3.75%) |
| 18.5 – 24.9 | 36 (45%) |
| 25 – 29.9 | 7 (8.75%) |
| ≥30 | 34 (42.5%) |
| Mean % body fat (SEM) | 34.6 (1.5) % |
| Mean % body water (SEM) | 47.2 (0.9) % |
| **Yale Food Addiction scale** |  |
| N participants without food addiction (%) | 67 (83.8%) |
| N participants with food addiction (%) | 13 (16.3%) |
| **HEI-2015** |  |
| Mean HEI-2015 (SEM) at baseline | 49.8 (1.8) |
| HEI-2015 range (0-100) | 25.2 – 72.2 |
| **Hunger ratings** |  |
| Mean hunger ratings at **baseline** (SEM) | 3.7 (0.2) range (1 to 7) |
| Mean hunger ratings **after MRI** (SEM) | 4.8 (0.2) range (1 – 7) * |
| Mean Total Physical activity (SEM) | 2020 (204) MET-min/week |

\*Hunger ratingBaseline vs hunger ratingafter MRI = (t(74) = -6.7, p< 0.001, Cohen's d = -0.77, paired, two-tailed t-test. BMI: Body Mass Index, HEI is the healthy eating Index, MET-min/week – metabolic equivalent, defined as the amount of oxygen consumed while sitting at rest and is equal to 3.5 ml O<sup>2</sup> per kg body weight x min. SEM – standard errors of the mean.

#### Motivational interviewing

The motivational interviewing session was divided into four phases: The interviewer, who was a dietitian trained in MI and is the first author of the study, elicited the participants to reflect on the following: (1) Their dietary patterns and whether there was a desire for a change, and (2) the advantages and disadvantages of the behavior they would like to change. The experimenter then asked participants to (3) rate their importance and readiness for change on a 10-point Likert scale and (4) verify whether they still would like to change an aspect of their dietary habits. The brief motivational intervention was audio-recorded, and five change and 5 sustain talk statements were extracted for use during an fMRI session at the second visit one week later. Phrases were chosen with the agreement of two different raters trained in MI.

#### Visit 2: Functional magnetic resonance imaging during value-based dietary decision-making

A week later, participants underwent an fMRI session after overnight fasting. Before the fMRI session, and to ensure a balanced distribution of healthy, unhealthy, tasty, and untasty food stimuli across the two talk conditions, participants rated each food item in terms of healthiness and tastiness on a 4-point Likert scale: strong no, no, yes, strong yes. Additionally, hunger ratings were measured on a 7-point Likert scale before and after the fMRI session. Importantly, these hunger ratings were averaged across the rating of three aspects of hunger: hedonic (i.e., How pleasant would it be to eat, now?), homeostatic (i.e., How much could you eat, now), and general (i.e., How hungry are you, now?) (Table 5).

#### Dietary decision-making task

To study the effect of change and sustain talk on dietary decision-making, participants performed an incentive compatible value-based dietary decision-making task.

The goal of the task was to rate snack food items of varying tastiness and healthiness on whether participants wanted to eat them for real at the end of the experiment, and after having listened to their change or sustain talk statements from the motivational interviewing session one week earlier.

As shown in Figure 5, the task counted ten blocks of 15 choice trials. Each block started with the instruction to consider the statement for change or sustaining eating habits. Then, one talk statement was played for 15 seconds, and the corresponding verbatim could be read on the screen as subtitle. For example, a sustain talk statement could be: "I won’t see an immediate effect on my weight. So, I tell myself: It’s not that bad, I can finish this bowl of chips". On the contrary, a corresponding change talk statement could read: "In terms of weight, I find that I have put on some weight recently, and I think it has to be that [referring to the bowl of chips]." Participants were asked to read and listen to their voices expressing the change or sustain talk statements, respectively.

**Figure 5.**
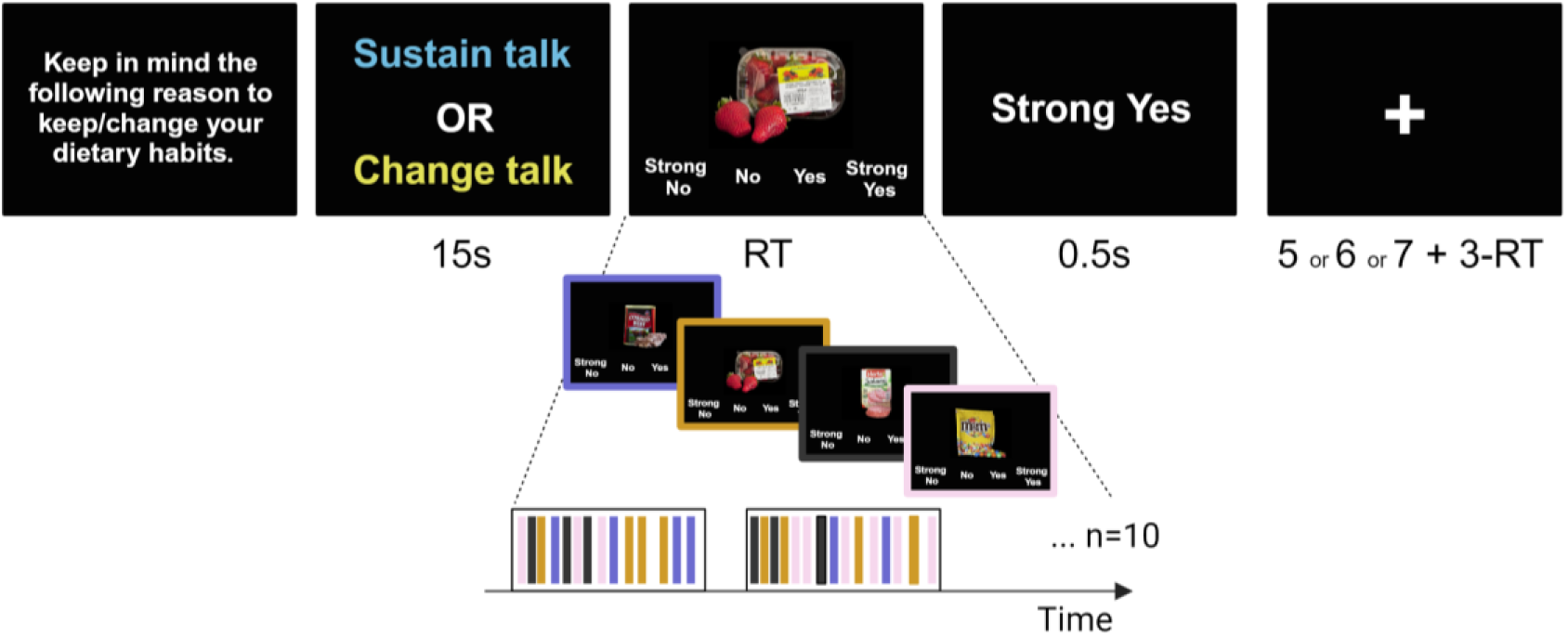
Value-based dietary decision-making task. Panels display screenshots of successive events within a change and sustain talk block. Each block started with a short reminder to consider the following statement to change/sustain current dietary habits. This information was followed by a change or sustain talk statement, which participants listened to and could also read on the screen. Following the statement, participants performed 15 food choices. Food items with varying levels of healthiness and tastiness were presented in a balanced way across all blocks. Each choice trial started with a food displayed on the screen, and participants had to rate on a 4-point Likert scale how much they wanted to eat the food for real at the end of the experiment. As soon as they hit a corresponding choice button, the choice was displayed on the screen. Trials within each talk block were separated by a fixation cross. Durations of each event within a trial are displayed in seconds. RT – reaction time.

After listening and reading the statements, participants rated 15 snack food stimuli on whether they wanted to eat this food for real at the end of the experiment. We refer to this rating as stimulus value. Participants used a 4-point Likert scale ranging from strong no to strong yes to indicate their choice (rating). Trials were separated by a jittered fixation cross (Figure 1).

#### Technical details of the value-based dietary decision-making task

In total the five change and five sustain talk blocks were randomly intermixed and counted 15 choice trials for a total of 75 food choice trials following a change talk statement and 75 food choice trials following a sustain talk statement. Change and sustain talk statements were selected from each participant’s motivational interviewing session that took place one week before the fMRI session.

Importantly, the task was performed in the fasted, hungry state (i.e., after overnight fasting, for hunger ratings see table 4), and it was incentive-compatible following similar procedures in the field of decision neuroscience (27, 31). To this aim, one food was chosen out of a chance for consumption at the end of the experiment. This guided the participants to adopt the optimal strategy of treating each decision as if it were the only one and hence to report their true preference for a given food stimulus. The foods that were eventually presented to the participant for consumption comprised chocolate bars, fruits (e.g., banana, clementine), or dairy-based drinks following previous work (25). Participants did not know what food they were going to be presented with during the experiment and only found out at the very end before leaving the laboratory.

A small break was scheduled inside the fMRI scanner after 75 food choice trials (i.e., 5 blocks), and a brief practice session occurred outside the MRI beforehand. Headphone volume was adjusted to each participant’s sensibility during a second practice session inside the MRI. The food pictures were presented on a computer screen as high-resolution images (72 dpi). MATLAB and Psychophysics Toolbox extensions (44) were used for stimulus presentation and response recording. Participants saw the food stimuli via a head-coil–based mirror and indicated their responses using an fMRI-compatible response box system. The hands used to make choices, and the display of the 4-point Likert scale (from left to right or right to left) was counterbalanced across participants.

The food stimuli were selected from a database of 600 food pictures used in previous studies (25, 26, 28). Prior to starting the dietary decision-making task and outside the MRI this study’s participants also rated each food stimulus on its healthiness and tastiness using a 4-Point Likert scale from strong no to strong yes. These ratings were used to (1) distribute tasty, untasty, healthy and unhealthy food stimuli in a balanced way across the two talk conditions, and (2) to test how tastiness and healthiness determined food wanting in the statistical analyses described below. Additionally, to check if participants were hungry, self-reported hunger was measured on a 7-point Likert scale (0, not hungry at all, 7=the hungriest one could imagine) both before and after the fMRI session. Importantly, these hunger ratings were averaged across the rating of three aspects of hunger: hedonic (i.e., How pleasant would it be to eat, now?), homeostatic (i.e., How much could you eat, now), and general (i.e., How hungry are you, now?).

#### Questionnaires and body composition measures

After the fMRI session, anthropometric measures such as body weight and body fat percentage were assessed by bio-impedance analysis using Tanita® Body Composition Analyzer SC-240MA (Tanita Corporation, Tokyo, Japan). Height was self-reported. The habitual physical activity level was estimated via the short form of the International Physical Activity Questionnaire (IPAQ-SF), which assessed the frequency and duration of physical activity according to intensity categories (moderate, vigorous, and walking) and sitting time over the previous seven days. Estimates of total physical activity are given in metabolic equivalent task-min/week (MET-min/week) (45). Participants filled out the Yale Food Addiction Scale (YFAS) to measure addictive-like eating habits (46).

#### Brain imaging data acquisition

Brain images were collected on a 3 T Verio Siemens scan using a 32-channel receive-only head coil. For the structural acquisition, whole-brain high-resolution T1-weighted structural scans were acquired with a magnetization-prepared rapid gradient echo (MPRAGE) sequence with the following parameters: TR = 2.4 s; TE = 2.05 ms; flip angle=9°, slice thickness=0.8mm. Multi-band T2*-weighted multi-echoplanar images (mEPI) with BOLD contrast were acquired for the functional acquisition. To cover the whole brain with a repetition time of 1.25 seconds, the following sequence parameters were applied: echo times 14.8 ms; 33.4 ms and 52 ms; 48 slices collected in an interleaved manner; 3-mm slice thickness, FOV = 192 mm; flip angle = 68°, and an average of 646 volumes per participate session. Three echos were acquired to accommodate better the compromise between spatial resolution and signal of the orbitofrontal context (47, 48), which comprises the ventromedial prefrontal cortex, a brain region of interest. To further minimize the signal dropout in the orbitofrontal cortex, an oblique acquisition orientation of 30° above the anterior-posterior commissure line was applied (49).

#### fMRI data pre-processing

Anatomical MPRAGE and functional mEPIs were processed using the Statistical Parametric Mapping – SPM12 (Welcome Department of Imaging Neuroscience, Institute of Neurology, London, United Kingdom) implemented in MATLAB version R2020b (The MathWorks Inc., United States). Each participant’s anatomical image was segmented into gray, white matter, and cerebrospinal fluid using the SPM12 segmentation tool. The three echo images of each volume were summed into one EPI volume using the SPM 12 Image Calculator (50-52). Then, the summed EPIs were spatially realigned and motion corrected, coregistered to the mean image, normalized to the Montreal Neurological Institute (MNI) space using the same transformation as the anatomical image, and spatially smoothed using a Gaussian kernel with a full-width-at-half-maximum of 8 mm.

#### Behavioral and brain data analyses

Statistical tests were conducted with the Matlab Statistical Toolbox (Matlab 2020a, MathWorks), and JASP (JASP 0.16.1).

#### Behavioral analyses

First, we tested whether change talk and sustain talk had different effects on overall food valuation and whether these effects were different in participants with normal BMI and participants with overweight and obese BMI. To this aim a mixed effects analyses of variance tested for a main effect of type of talk, BMI group (< 25 kg/m^2^ and ≥ 25 kg/m^2^) and the interaction type of talk by group. The model assumed random intercept nested by participant IDs and used Likelihood ratio tests. Paired, two-tailed t-tests were used to compare the mean stimulus value ratings and reaction times between BMI groups and type of talks. Pearson’s correlational analyses tested if food preferences and reaction times were moderated by participant’s BMI and the number of symptoms on Yale Food Addiction Scale (YFAS).

To test how tastiness and healthiness ratings were considered during dietary decision making the mean-centered stimulus values (SV, coded -2 for "strong no," -1 for "no," 1 for "yes," and 2 for "strong yes"), were fit to a multilevel general linear model (GLM1). GLM 1 included on the individual first level the following regressors: trial number, to account for fatigue effects, healthiness ratings, tastiness ratings, two categorical indicators of sustain and change talk blocks. Additionally, GLM1 included four interaction terms: STxHR, CTxHR, STxTR, and CTxTR. These interactions tested for effects of healthiness and tastiness on stimulus value in trials following change and sustain talk, respectively. The individual beta coefficients for these interaction regressors were fit into a second-level factorial analysis of variance (ANOVA) tested for main effects of talk (change versus sustain talk), food attribute (healthiness versus tastiness rating), and the interaction type of talk by type of food attribute. Post hoc, paired, two-tailed t-tests were used to determine the directionality of interactions.

To investigate the effects of type of talk as a function of continuous BMI and food addiction score regulatory success scores were calculated for each participant and for choice trials following change and sustain talk, respectively. The regulatory success score was computed for each talk condition by subtracting the individual beta coefficients for tastiness and healthiness on stimulus value from GLM 1 :

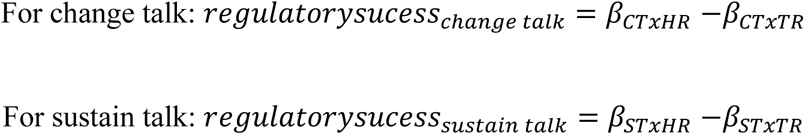

Positive values meant that healthiness determined food stimulus values and hence wanting more than tastiness. Negative regulatory success scores meant that the tastiness determined food wanting more than healthiness. A mixed effects ANOVA then tested regulatory success for main effects of type of talk, BMI group and food addiction scores (i.e., YFAS number of food addiction-like symptoms) and all possible interactions using Likelihood ratio tests and considering random intercepts nested by participant number.

To unpack detected interactions Pearson’s correlations were conducted between regulatory success scores and food addiction scores in change talk and sustain talk conditions and across participants with normal BMI and participants with overweight and obesity.

#### Brain imaging statistical analyses

fMRI timeseries were fitted using multilevel general linear models (GLMs). To localize brain regions encoding food stimulus values irrespective of talk GLM2 included on the first level the following regressors: (1) an onset regressor at time of talk with boxcar durations of 15 seconds, (2) a choice onset regressor with a boxcar duration corresponding to individual reaction times, (3) a parametric modulator at time of choice consisting of stimulus value ratings, and (4) missed trials indicator with a boxcar duration of 3 seconds.

A third GLM3 was used to localize brain activation associated to stimulus value encoding following change compared to sustain talks. GLM3 was analogous to GLM2, but split down the choice onsets and stimulus value parametric regressions into whether they followed sustain and change talks respectively. In more detail the first level six regressors of interest were: (1) an indicator for talk display with boxcar durations of 15 seconds, (2) choice onset regressors at time of food image display following change talk with durations corresponding to the reaction time, (3) a parametric modulator at time of choice consisting of stimulus value after change talk, (4) choice onset regressors at time of food image display following sustain talk with durations corresponding to the reaction time, (5) a parametric modulator at time of choice consisting of stimulus value after sustain talk, and (6) missed trials with a boxcar duration of 3 seconds.

Six realignment parameters (x, y, z, roll, pitch, and yaw) were added as regressors of non-interest to correct for head movement. Boxcar functions for each trial were convolved with the canonical hemodynamic response function in SPM. Individual contrast images for onset choice and the parametric modulator, the stimulus value, were then fitted into a second-level random effects analysis that used one-sampled t-tests and paired t-tests to compare change to sustain talks conditions. BMI was added as covariate in the second-level random effects analysis to GLM 2. Note, GLM2 was also used to define the seed region for the PPI analysis and which was located within the vmPFC in response to stimulus value ratings.

#### Psycho-physiological-interaction (PPI) analysis

For the PPI analysis we used a GLM 4 like GLM 2, where choice onset was modeled as a single choice onset regressor (duration = reaction times), but parametric modulations by stimulus values were removed. The model further controlled for variance explained by talk display (duration = 15 s), missed trials (duration = 3 s) and head movement by including the six realignment parameters as regressors of non-interest. BOLD signals from a 4-mm radius sphere were extracted from a seed region located in the vmPFC and centered on the MNI coordinates that correlated significantly to stimulus value in GLM 2 (MNI = [0, 38, -6], Table 2)). Choice onset following change talk and sustain talks were respectively modeled trial-by-trial as 3 seconds boxcars. The vmPFC signal was deconvolved and multiplied with the psychological contrast of choice onset after change talk minus sustain talk to obtain the interaction term. The three regressors (i.e., physiological vmPFC signal, psychological choice onset contrast change > sustain talk and the psycho-physiological interaction) were then convolved with the canonical hemodynamic response function and entered into a first-level PPI-GLM for each participant. Individual betas for the PPI regressor were fit into a second-level two-sample t-test where participants with BMI<25 were compared to participants whose BMI≥25 using two-sampled t-tests. In addition, beta estimates from the PPI regressor were extracted from a 5 mm radius sphere centered around the peak coordinate located in the dlPFC (MNI = [22, 58, 6]).

#### Responses of the neurobiological craving signature (NCS) to food stimulus value (i.e., choices)

To assess how change and sustain talk altered craving-related brain responses (as measured by the Neurobiological Craving Signature following Koban et al. (2023) (26), a multilevel GLM 5 was fit to each participant’s timeseries at the first level. GLM 5 included ten regressors of interest: (1) an indicator for talk display and listening, (2) an onset regressor for tasty food choices following change talk, (3) stimulus value as a parametric modulator of the tasty food choices following change talk, (4) an onset regressor for healthy food choices following change talk, (5) stimulus value as a parametric modulator of the healthy food choices following change talk, (6) an onset regressor for tasty food choices following sustain talk, (7) stimulus value as a parametric modulator of the tasty food choices following sustain talk, (8) an onset regressor for healthy food choices following sustain talk, (9) stimulus value as a parametric modulator of the healthy food choices following sustain talk, and (10) onset regressors for missed trials with boxcar durations of 3 seconds. The six realignment parameters were included as regressors of non-interest to control for head movement.

The contrasts of interest were the parametric regressors (3), and (7) predicting BOLD responses at the time of food choice in correlation to stimulus values (i.e. wanting ratings) for tasty foods after change and sustain talk. Note, tasty foods could be both healthy and unhealthy, and healthy foods could be both tasty and untasty.

The Neurobiological Craving Signature responses were quantified as the dot product between these contrast images of interest for each participant and the Neurobiological Craving Signature map, plus a fixed intercept. The NCS consists of a map of beta values assigned to each voxel of the brain that reflected their weight in predicting food wanting (cravings) in Koban et al. (2023). This yielded one value per participant and contrast of interest, which reflected how much the Neurobiological Craving Signature responded to tasty and healthy foods in the different self-regulation conditions.

Individual dot product values were then entered into a second-level random effects analysis, which used a mixed effects ANOVA to test for main effects of type of talk (change versus sustain talk), type of food (tasty versus healthy) and their interaction. The model further controlled for variance potentially explained by food addiction (i.e., number of food addiction-like symptoms on the YFAS) on the fixed effects level. It assumed random slopes for the effects of talk, type of food, and a random intercept, all nested by participant number, and used likelihood ratio tests. To further explore the relationship between BMI group (participants with normal to overweight versus participants with obesity) and NCS responses, Pearson’s correlation analyses were conducted between BMI and NCS responses to tasty and healthy foods following change talk and for each BMI group, respectively.

#### Definition of regions of interest

The following regions of interest were used:

1. The vmPFC as a seed region for PPI analyses was defined by the MNI coordinates [0, 38, -6], which activated significantly in response to stimulus value (i.e., food wanting ratings) at time of food choices (pFWE<0.05). Eigenvalues from a 5-mm sphere centered around these peak coordinates were extracted for the physiological regressor and multiplied with the contrast choice onsets following change versus sustain talk to form the PPI regressor.
2. The dlPFC region of interest for small volume correction was defined by the MNI coordinates [-48 15 24], which were reported by Hare et al. Science 2009 for areas activating stronger in successful self-controllers during dietary decision-making.

## Study-pre-registration

This study was pre-registered (https://clinicaltrials.gov/study/NCT05101863). The authors declare no competing interest.

## One Sentence Summary

Pondering behavior change affects food preferences and the brain’s valuation and cognitive control systems in persons with obesity and food addiction.

## Data availability

Raw behavioral data cannot be shared due to GPDR requirements. Metadata for figures are shared on the study’s OSF website: https://osf.io/4adr3/.

## Code availability

Code to analyze behavioral data and food decision-making task scripts are available on the study’s OSF website (https://osf.io/4adr3/). The NCS and code to apply it to any fMRI data (apply_ncs.m function) are publicly available on Github (https://github.com/canlab/Neuroimaging_Pattern_Masks/tree/master/Multivariate_signature_patterns/2022_Koban_NCS_Craving).

## Acknowledgments

We thank Hilke Plassmann and Nathalie George for their insightful discussions at various steps of the project. We thank the CENIR team for their support and collaboration throughout the project, and Valentine Lemoine (clinical research assistant, ICAN) and the ICAN platform for help in recruitment of the participants with obesity.

## Authors Contributions

BR and LS conceived and designed the study with advice from CP, JMO, PF, and JYR. BR, MR, IK, SF, JYR and CP included participants. BR collected data with help from MR, BF, SF, and IK. BR, MR. BR and HA analyzed the data under the supervision of LS and advice from LK. BR and LS wrote a first draft of the manuscript, and all authors contributed to the final text.

## Supplementary material

**Figure S1.**
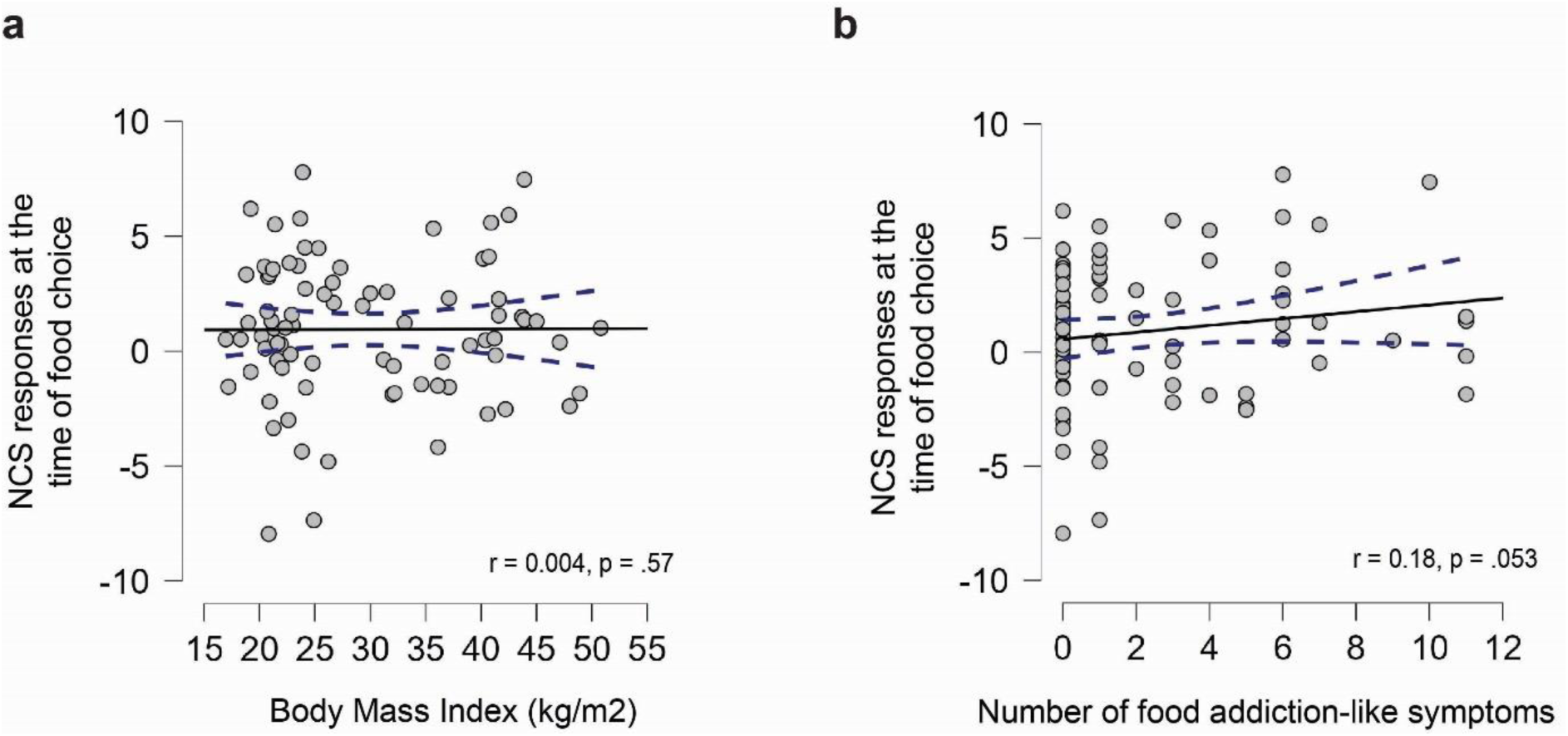
NCS responses at the time of food choice. Correlation between the Neurobiological Craving Signature (NCS) responses at the time of food choice with **(a)** body mass index (Pearson’s r = 0.004, p = 0.57, n=80, one-sided) and **(b)** particle correlation of the NCS responses at the time of food choice and number of food addiction-like symptoms (Pearson’s r = 0.18, p = 0.053, n=80, one-sided, BMI partialled out). Each dot corresponds to a participant; the blue lines designate 95% confidence intervals.

## Methods

### Dietary intake assessment

To ascertain the required nutrition-related variables, once the 24h-recall was collected, energy intake (in Kcal), sodium and fatty acids quantities were ascertained through Nutrilog SAS software (Nutrilog-Online 20231010). Additionally, to measure diet quality, the healthy eating index (HEI) was computed (57). The HEI-2015 is based on the Dietary Guidelines for Americans 2015-2020 (for a comparison between the American guidelines and national guidelines see SI Appendix Table S4). It has 13 items whose scoring follows adequacy- or moderation-based energy-adjusted intakes cut-offs, except for the fatty acids components. Each components weights differently into the final scoring (0-100, where higher scores indicate a healthier dietary pattern): total fruits (5 points), whole fruits (5 points), total vegetables (5 points), greens and beans (5 points), whole grains (10 points), dairy (10 points), total <u>protein foods</u> (5 points), fatty acids (polyunsaturated fatty acid plus <u>monounsaturated fatty acid</u> to <u>saturated fatty acid</u> ratio; (10 points), refined grains (10 points), sodium (10 points), added sugars (10 points), and saturated fats (10 points). Refined grains, sodium, added sugars, and saturated fats as components to be consumed in moderation have a reversed scoring (Krebs-Smith et al., 2018). The HEI score for the sample is shown in Table 5.

