## Supplementary Material for "The neural pathways of change: An fMRI study of the effects of behavioral change suggestions on value-based dietary decision-making"

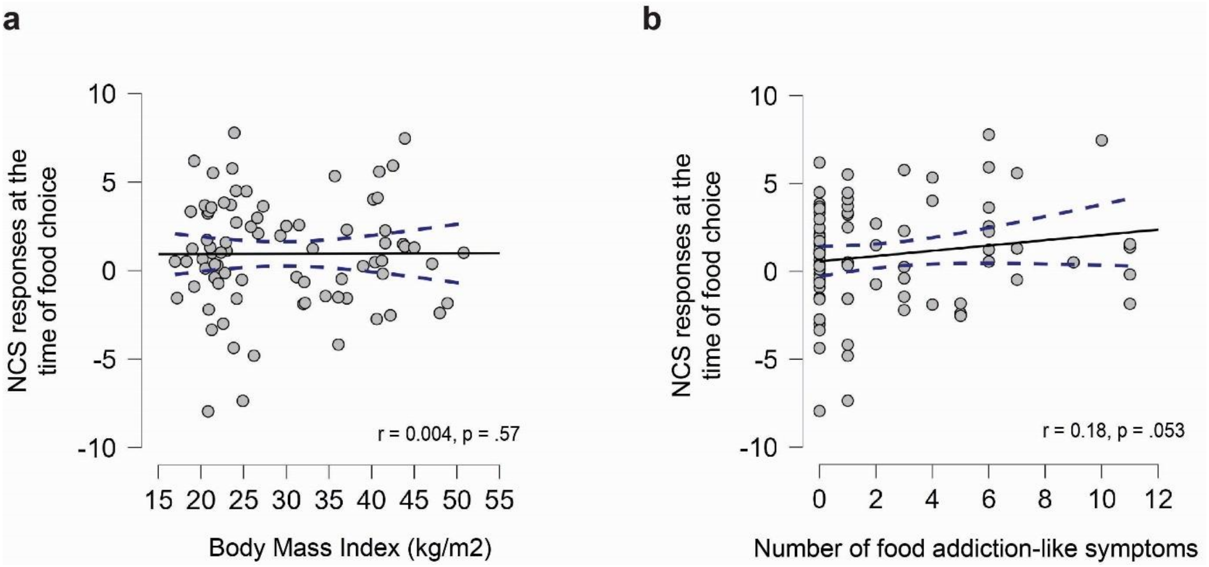


**Figure S1**. **NCS responses at the time of food choice.** Correlation between the Neurobiological Craving Signature (NCS) responses at the time of food choice with **(a)** body mass index (Pearson's r = 0.004, p = 0.57, n=80, one-sided) and **(b)** particle correlation of the NCS responses at the time of food choice and number of food addiction-like symptoms (Pearson's r = 0.18, p = 0.053, n=80, one-sided, BMI partialled out). Each dot corresponds to a participant; the blue lines designate 95% confidence intervals.

### Methods

#### Dietary intake assessment

To ascertain the required nutrition-related variables, once the 24h-recall was collected, energy intake (in Kcal), sodium and fatty acids quantities were ascertained through Nutrilog SAS software (Nutrilog-Online 20231010). Additionally, to measure diet quality, the healthy eating index (HEI) was computed (57). The HEI-2015 is based on the Dietary Guidelines for Americans 2015-2020 (for a comparison between the American guidelines and national guidelines see SI Appendix Table S4). It has 13 items whose scoring follows adequacy- or moderation-based energy-adjusted intakes cut-offs, except for the fatty acids components. Each components weights differently into the final scoring (0-100, where higher scores indicate a healthier dietary pattern): total fruits (5 points), whole fruits (5 points), total vegetables (5 points), greens and beans (5 points), whole grains (10 points), dairy (10 points), total protein foods (5 points), fatty acids (polyunsaturated fatty acid plus monounsaturated fatty acid to saturated fatty acid ratio; (10 points), refined grains (10 points), sodium (10 points), added sugars (10 points), and saturated fats (10 points). Refined grains, sodium, added sugars, and saturated fats as components to be consumed in moderation have a reversed scoring (Krebs-Smith et al., 2018). The HEI score for the sample is shown in Table 5.
